# DeepCOVID: An Operational Deep Learning-driven Framework for Explainable Real-time COVID-19 Forecasting

**DOI:** 10.1101/2020.09.28.20203109

**Authors:** Alexander Rodríguez, Anika Tabassum, Jiaming Cui, Jiajia Xie, Javen Ho, Pulak Agarwal, Bijaya Adhikari, B. Aditya Prakash

## Abstract

How do we forecast an emerging pandemic in real time in a purely data-driven manner? How to leverage rich heterogeneous data based on various signals such as mobility, testing, and/or disease exposure for forecasting? How to handle noisy data and generate uncertainties in the forecast? In this paper, we present DeepCovid, an operational deep learning framework designed for real-time COVID-19 forecasting. Deep-Covid works well with sparse data and can handle noisy heterogeneous data signals by propagating the uncertainty from the data in a principled manner resulting in meaningful uncertainties in the forecast. The deployed framework also consists of modules for both real-time and retrospective exploratory analysis to enable interpretation of the forecasts. Results from real-time predictions (featured on the CDC website and FiveThirtyEight.com) since April 2020 indicates that our approach is competitive among the methods in the COVID-19 Forecast Hub, especially for short-term predictions.

## 1 Introduction

### Motivation

The devastating impact of the currently unfolding global COVID-19 pandemic has sharply illustrated our enormous vulnerability to emerging infectious diseases. Forecasting disease trajectories is a non-trivial and important task. Estimating various measures related to the epidemic gives policymakers valuable lead time to plan interventions and optimize supply chain decisions. Hence, accurate forecasts are critical in combating epidemic outbreaks including the current pandemic (Holmdahl and Buckee 2020).

To encourage research in epidemic forecasting and to provide a unified prediction platform, the US Centers for Disease Control and Prevention (CDC) organized a collaborative forecasting task under the umbrella of the COVID-19 Forecast Hub (Reich et al. 2020). The forecasting targets include various COVID-related metrics including mortality and hospitalizations at various temporal and spatial resolutions. The initiative has attracted submissions from more than 58 teams from industry and academia (as of Sept 2020).

Majority of the participating approaches can be classified into two categories (i) mechanistic & (ii) statistical. Mechanistic approaches model the underlying disease transmission over the population under various assumptions like using ordinary differential equations (Zhang et al. 2017) and/or agent based models (Venkatramanan et al. 2018). While valuable for long-term ‘what-if’ scenario generation, mechanistic approaches have several challenges in real-time forecasting of an emerging pandemic. The first drawback is that the complexity of the model rises with the number of data sources being used. Hence, to manage the complexity, most models often use only one or two data signals (such as weather) along with the observed disease incidence (Shaman, Goldstein, and Lipsitch 2010). Hence it is not trivial to extend these models to include various other data signals (e.g. social media data).

On the other hand, statistical approaches are fairly new in epidemic forecasting. They exploit correlations between various data sources and the forecast targets to learn a functional dependence between the two and use the learned function to make predictions (Yuan et al. 2013). There are several challenges in designing statistical approaches, as we discuss later (see Sec. 4). Most of the existing statistical approaches for epidemic forecasting (usually developed for influenza forecasting, including work by the authors (Adhikari et al. 2019)) cannot be readily adapted for COVID forecasting due to several issues such as lack of historical data and poor data quality. Indeed, to our best knowledge, there is no work in the literature describing a completely data-driven approach for real-time forecasting.

### Our goals

In this paper, we describe our deployed frame-work DeepCovid (see Fig.1), the *first* purely data-driven deep learning (DL) model for real time pandemic forecasting in the COVID-19 Forecast Hub (Reich et al. 2020)^1^ and the official ensemble. Our real-time forecasts are showcased in the hub, the CDC website^2^ and the popular FiveThirtyEight website^3^. By our work, we aim to address a gap in the literature pertaining to using purely data driven approaches for emerging pandemics, with the following four goals. *(G1) Coping with heterogeneous, scarce and noisy data:* A DL-based model can ingest many heterogeneous signals that are more sensitive to what is happening on the ground, without laborious feature engineering. To fully take advantage of this, our framework is designed with careful consideration of the data and modeling challenges faced in robust real-time forecasting with principled uncertainty estimation. *(G2) Bring a complementary forecasting perspective:* A good ensemble needs diverse perspectives (Reich et al. 2019; Ray et al. 2020).The overwhelming majority of the teams in the ensemble perform mechanistic modeling. By being the first data-driven DL method in the ensemble, DeepCovid brings a unique perspective (Sheridan 2020) closer to the observed data signals with minimal assumptions. *(G3) Accurate short-term forecasting:* The utility of statistical models in short-term forecasting has been observed before (Holmdahl and Buckee 2020), which are useful to plan intervention and allocate resources. We demonstrate that DeepCovid excels in this task by comparing to a strong baseline. *(G4) Enable communication and interpretation:* This is to ensure our framework gives explanations for its forecasts, which are very important for communication and interpretation by both the public and decision makers.

**Figure 1:**
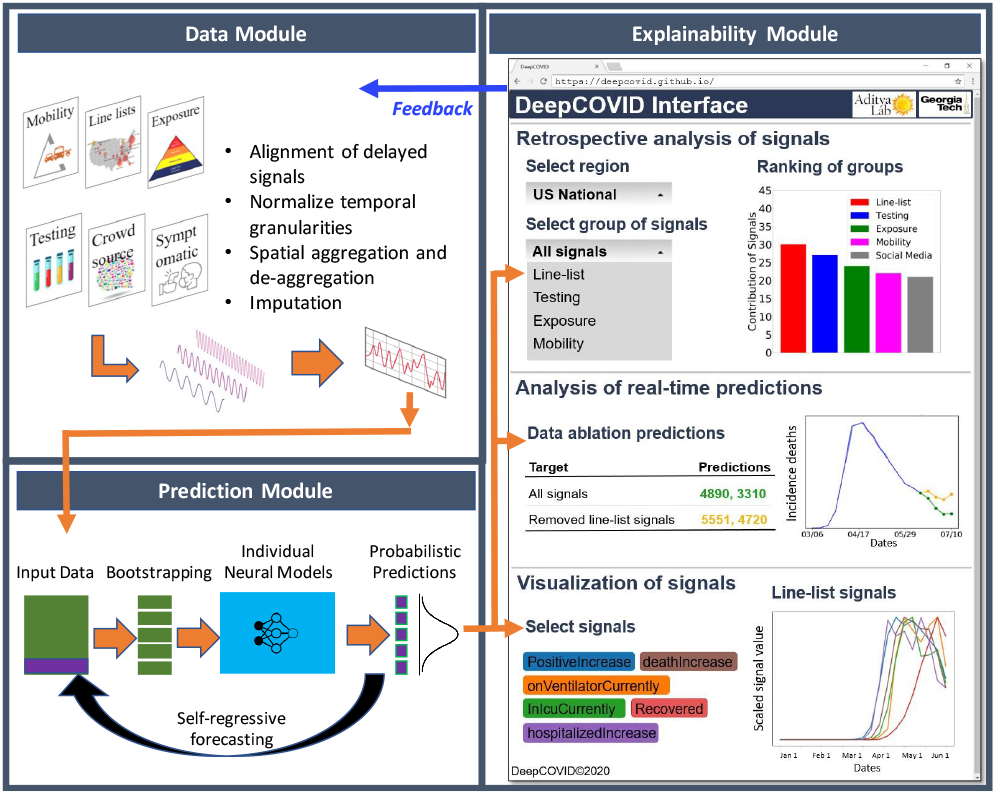
Schematic of our DeepCovid framework for real-time COVID-19 forecasting. The data module is dedicated to data pre-processing including imputation of missing values and aggregating at the right temporal and spatial resolution. The prediction module generates probabilistic forecasts based on the curated data. Finally, the explainability module (with interface) allows both the real-time and retrospective analysis of forecasts to build intuitive explanations.

### Contributions

Our contributions are as follows:

- We propose DeepCovid, one of the first deep learning based real-time COVID forecasting frameworks, whose performance in the CDC COVID-19 Forecast Hub (since April 2020) demonstrates that it is consistently competitive, especially at short term forecasting.
- Using the current pandemic as a testbed, we address several challenges of such real time forecasting including interpretability and uncertainty estimation in a principled fashion.
- We provide valuable observations and ‘lessons learned’ from our experience for modeling emerging infectious diseases using purely data-driven methods.

The rest of the paper is organized in the following way: we give additional related work and background next, then describe the modules in our framework, then empirical results and finally conclude with discussion. Appendix and other resources can be found online^4^.

## 2 Related Work

### Epidemic Forecasting

Modeling approaches for epidemic forecasting can be broadly categorized into mechanistic (Zhang et al. 2017; Shaman and Karspeck 2012) and statistical (Tizzoni et al. 2012; Osthus et al. 2019; Brooks et al. 2018), the latter closely related to time series analysis (Box et al. 2015; Jha et al. 2015). Past work, especially in the context of influenza, has explored leveraging multiple sources of data, from search engine (Ginsberg et al. 2009; Yuan et al. 2013) to weather data (Shaman, Goldstein, and Lipsitch 2010; Tamerius et al. 2013; Volkova et al. 2017). Deep learning for epidemic forecasting is gaining more research interest lately. Recent works include (Adhikari et al. 2019), which learns low-dimensional embeddings of influenza seasons for forecasting and (Wang, Chen, and Marathe 2019) which exploits both intra and inter seasonal similarity of historical seasons.

### COVID-19 Forecasting

Other approaches adopted by the contributing models are mechanistic (Zou et al. 2020; Chinazzi et al. 2020; Baek et al. 2020) and statistical (Altieri et al. 2020; Murray et al. 2020). The official hub ensemble (Ray et al. 2020) combines forecasts from ours and other models.

## 3 Background

### Forecasting requirements from CDC

Starting in April 2020, CDC requests probabilistic forecasts for COVID-19 associated mortality and hospitalizations at various temporal and spatial resolutions to be used by policymakers to plan intervention and allocate resources. As mentioned before, our model has been part of the ensemble since its inception.

### Targets

The forecasting targets are the following: *(T1) Incidence and cumulative weekly deaths:* Reported incidence (new) and cumulative deaths for US states and the US overall. The data reported by Johns Hopkins University (JHU) (Dong et al. 2020) serves as gold standard for the CDC. *(T2) Incidence daily hospitalizations:* Reported new hospitalizations for US states and the US overall. CDC had not fixed a gold standard for this (as of Sept 2020) but we found the data provided by the COVID Tracking Project^5^ to be the most complete.

### Problem formulation

We can state our real-time forecasting problem for a specific geography as follows.

### Given

an observed multivariate time series of COVID-related signals 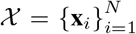 and corresponding values for the forecasting target 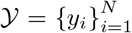, where *N* is the size of the sequence until the current date.

### Predict

next *k* values of forecasting target, i.e.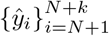, where *k* = 4 for T1 (4 weeks ahead) and *k* = 28 for T2 (28 days ahead).

## 4 Our Framework: DeepCOVID

We built DeepCovid, our framework for explainable real-time COVID-19 forecasting, which contains three modules depicted in Fig. 1: data module, prediction module, and explainability module. By separating data and prediction modules, our goal is to differentiate between the handling of noisy data from the learning process. For the predictive module, we use deep learning because it is a flexible, scalable, and efficient technology, and an excellent choice to model non-linearities, with the capability to ingest heterogeneous datasets. As mentioned before, explainability is a challenge in data-driven models. We want to understand and connect forecasts with epidemiological reasons. Once we have insights about our predictions, we will have a feedback loop to improve performance. Therefore, we have an explicit module for explainability which helps to shed some light on our predictions in a dynamic situation. We next discuss challenges and our approaches for our modules in more detail (see Table 1 for a summary).

**Table 1:** Summary of challenges and solutions

| Aspect | Challenges | Our Approaches |
| --- | --- | --- |
| Data | (C1) *Data collection*: Data signals come from multiple sources with formats varying over time | Create a flexible data extraction pipeline specialized to each data format |
|  | (C2) *Selection of signals*: Data signals need to represent the underlying disease spread and enable insights and interpretation | Select epidemiologically meaningful data, connecting to mechanistic insights, which can represent underlying dynamics, syndromic states, exposure, connectivity etc. Also use explainability module to filter out ineffective signals |
|  | (C3) *Temporal misalignment*: Signals have various reporting delays, temporary pauses in reporting, and differ in temporal granularity | Handle delays by creating aligned versions of training data; for smaller reporting pauses treat as delays; try to normalize and convert granularities in a reversible way (daily to weekly and vice versa) |
|  | (C4) *Spatial misalignment*: Signals are reported at different spatial granularities (county vs state vs national etc) | Aggregate and de-aggregate signals considering their semantics |
|  | (C5) *Data quality and missing values*: Some predictors and even targets are missing, noisy, and unreliable for some geographical regions | Impute targets based on epidemiologically-grounded assumptions; tried learning dependence of features on each other; remove predictors when not available; also create easy-to-update visualizations to help assess their quality and utility efficiently in a dynamic way |
| Prediction | (C6) *Data Sparsity*: Scarce and limited data due to the novel and dynamic nature of the disease. Learning generalizable models in this setting is challenging. | Employ a neural architecture with a smaller number of parameters to avoid overfitting, and handle other inconsistencies separately |
|  | (C7) *Robust point and probabilistic forecasting*: Predictions need to be robust to noise and data issues; CDC requires predictions to always be accompanied by uncertainty bounds | Robust initialization of optimization; leverage bootstrapping for flexible and cheap quantification of epistemic uncertainty |
|  | (C8) *Temporal consistency between consecutive forecasts*: Each forecast further into the future should be consistent (both in values and uncertainties) with previous ones. Due to data sparsity we can not train architectures that could enforce this automatically (e.g. LSTM). | Use self-regressive forecasting - predictions of week k as part of training data for week k+1; this way we can also propagate uncertainty to future predictions in a principled way via sampling |
| Explainability | (C9) *Real-time insights of forecasts for decision-making and communication*: Decision-makers and public prefer forecasts relatable to epidemiological knowledge and allowing reasoning on their reliability. | Data ablation for all sets of signals for the *current* week to observe the predictive contribution of each set; use statistical significance tests to identify sets of signals driving the predictive behavior; visualize these important signals to reason about the reliability of such predictions |
|  | (C10) *Retrospectively understand signal strengths*: Signal strengths inform policy development and allow continual improvement of forecasts. | Data ablation for past predictions; use significance tests to identify set of signals affecting performance compared to ground truth; visualize their contribution and inform the selection of signals of the data module by removing the ones with negative contribution |

### 4.1 Data Module

In this section, we describe our data collection and preprocessing for input to our prediction module.

#### Challenges

Data pre-processing for real-time COVID forecasting brings several challenges primarily due to a novel emerging scenario. *(C1) Data collection:* Collecting data in such a chaotic scenario is challenging because it comes from multiple sources (often collected by volunteers), is in different formats, some even changing over time (e.g. reported deaths from JHU). *(C2) Selection of signals:* Selected signals should describe the different facets of the disease spread. To enable epidemiological observations about our predictions, this selection has to be driven by appropriate rationale. *(C3) Temporal misalignment:* Since the signals are collected from multiple sources, they often have temporal misalignment. Some signals presented 1-2 weeks of lag due to delays in reporting from hospitals, public records and government officials; furthermore, some temporary pauses in reporting occurred without previous notice. In addition to that, some signals have different temporal resolutions (days vs weeks). *(C4) Spatial misalignment:* Some records are reported at specific spatial granularity (county or state level) and their conversion to higher geographical levels is non-trivial. *(C5) Data quality and missing values* Most prominently, one of our forecasting targets, incident hospitalizations, has not been reported in 11 states (CA, DC, TX, IL, LA, PA, MI, MO, NC, NV, DE). Additionally, some predictor signals have been reported only in a few states (e.g. CDC-reported hospitalization rates).

#### Our approach

To address *(C1)*, we developed a flexible data extraction pipeline personalized for each data format to convert them all to a standard format. For *(C2)*, we extensively searched for meaningful signals from an epidemiological perspective. The outcome of our search is summarized in Table 2 for the signals consistently used in our predictions. We collected the signals for 52 geographical regions: US National, the 50 US states and Washington D.C. We address the data misalignment in *(C3)* as follows. For the lags in reporting, we downshift the signals. Our idea is to align all the signals based on their latest records since it is safe to assume that the latest records are more indicative for future targets. For smaller pauses in reporting, we treat them as delays. To address weekly/daily inconsistency, signals require different treatment depending on their nature. For example, since weekly hospitalization rate and CLI% ER visits have been recorded as percentages, we choose to consider the same value for daily incidences because it is not meaningful to transform it. For *(C4)*, signal 13 in Table 2 is recorded only at county-level, while other signals (6 and 7) contain records at the HHS region-level^6^. To provide a state-wise forecast model, we transformed the signal record state-wise by aggregating and de-aggregating, respectively. For this, it is crucial to consider the population of such geographic regions. Lastly, it is important to address *(C5)* in a meaningful way. For 11 states with missing values, we had related signals such as hospitalization and ICU patients. We inferred missing values by leveraging these signals and making reasonable assumptions (such as the effect of people’s recovery and death in one week). More details about handling these challenges and our data signals can be found in the appendix.

**Table 2:** Overview of data signals used in DeepCovid. (ILI=Influenza like Illness; CLI=COVID like Illness). Expanded table can be found in the appendix.

| Type/Rationale | Signals |
| --- | --- |
| (DS1) *Line list*: Traditional surveillance for tracking patients and symptoms | 1. Confirmed cases; 2. UCI beds currently occupied; 3. People on ventilation; 4. Recovered; 5. Hospitalization rate (COVID-Net); 6. ILI% ER visits; 7. CLI% ER visits; 8. Excess Deaths; |
| (DS2) *Testing*: Capture changing screening artifacts | 9. People tested; 10. Negative cases; 11. Emergency facilities reporting; 12. Number of providers; |
| (DS3) *Crowdsourced*: Symptomatic surveillance | 13. Digital thermometer readings provide ILI%; |
| (DS4) *Mobility*: Evidence of changing contact patterns | 14. Retail and recreation; 15. Grocery and pharmacy; 16. Parks; 17. Transit stations; 18. Residential; 19. Workplaces; 20. Overall-region-based |
| (DS5) *Exposure*: Measure social contacts | 21-22. Device exposures (normal & adjusted); |
| (DS6) *Social Surveys*: Measure symptomatic burden | 23. CLI%; 24. ILI% |

### 4.2 Prediction Module

#### Challenges

Real-time COVID forecasting is a difficult problem with challenges originating from data and CDC requirements. Some of the challenges we encountered in designing the prediction module are the following. *(C6) Data Sparsity:* One of the major challenges in forecasting emerging disease,especially at an early stage, is the scarce and limited data due to the novel and dynamic nature of the disease. Extracting enough information from the few available data points to ensure generalizable forecasts is a challenging problem. *(C7) Robust point and probabilistic forecasting:* Forecasting disease in real-time is already challenging, and the problem is even more difficult in presence of data issues mentioned above and public health requirements (CDC requires predictions to always be accompanied by uncertainty bounds). Hence, the questions we tackle in designing our framework are (1) how do we principally quantify uncertainty of our forecasts?; and (2) how do we ensure that our approach is robust to the noise and other data issues such that ensures reliable point and probabilistic forecasts? *(C8) Temporal consistency between consecutive forecasts:* Due to data sparsity we cannot hope to train deep networks enforcing temporal consistency (values and uncertainties consistent with previous ones) such as LSTM and GRU. So the question is, how to design a neural architecture, which has few enough parameters to train from sparse data while ensuring temporal consistency between the forecasts?

#### Our approach

See Alg. 1. When we started to participate in the CDC task (April 2020), there was only 1 month of meaningful (non-zero) data; so it was unclear how to effectively train a recurrent neural network. Hence we opted to use a feedforward network with autoregressive inputs to incorporate short-term dependencies in the time series. To fully address *(C6)*, we also need to avoid overfitting, for which we empirically evaluated several parameters including the number of layers & sizes. We found that our network needed a small number of parameters to generalize well, specifically the following. Input layer size: number of signals to be included (may vary by period and geography depending on insights from explainability module) plus 1 for the target (i.e. one autoregressive input). Hidden layers size: We found that three hidden layers with sizes 10, 5 and 2 generalized well. We used ReLU activation function, which is especially useful in the output layer as we want our predictions to be positive.

##### Algorithm 1 Predictive module training

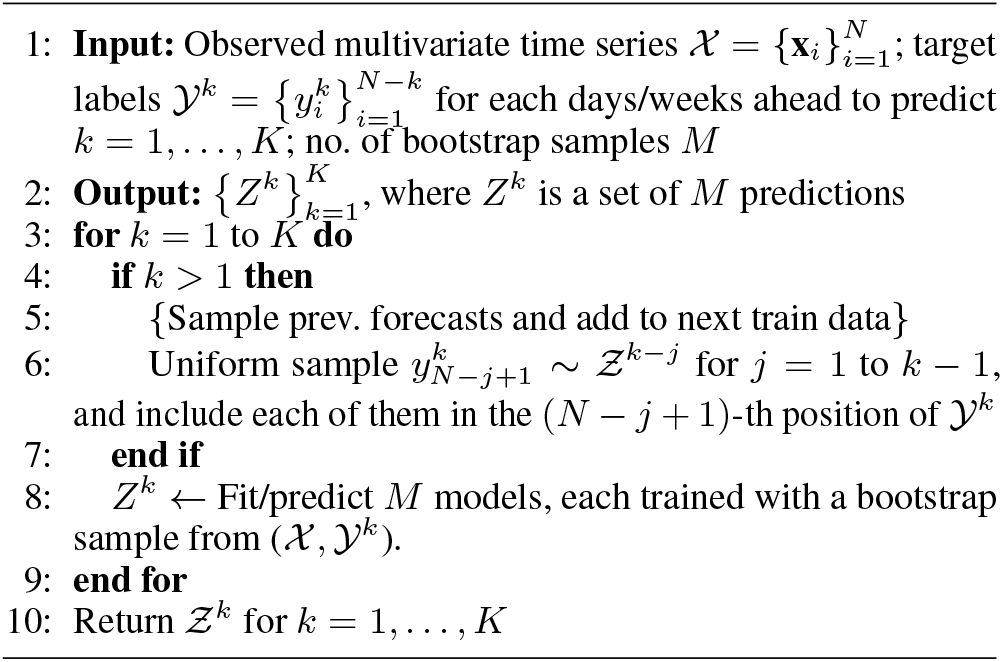

To address *(C7)*, we want to capture epistemic uncertainty coming from data in a principled way. Note that we want data-based uncertainty, not model-based uncertainty (e.g. MC dropout). Thus, we use bootstrapping (Gelman and Vehtari 2020): for each future incidence target *k* (lines 3-9), we generate one prediction per bootstrap sample so that we obtain a set of *M* predictions representing uncertainty in data (line 8)–note this process is ‘embarrassingly’ parallelizable. Robustness of our point and probabilistic predictions are closely related to optimization. Optimizing the parameters of a neural network with sparse, noisy and heterogeneous data is challenging. In fact, we found that our optimization sometimes was trapped in a local optima. Hence, to improve robustness, we used batch normalization (Ioffe and Szegedy 2015) to alleviate initialization problems. Beside that, for each bootstrap sample, we run the same optimization with different initializations and select the one that leads to lowest loss in the (training) data. We found that setting *M* = 80 and 20 initializations led to robust predictions in a timely manner with our computing resources.

To address *(C8)*, we enforce temporal consistency and capture temporal correlations between consecutive forecasts by using predictions from targets 1, …, *k*− 1 as part of the training data for target *k* (self-regressive forecasting, as noted in Fig. 1). In this way, we are also propagating uncertainty to future predictions (lines 4-7). This process can be regarded as a semi-supervised learning procedure because we use our model to create new labels that are used in training data for future targets.

*Note*: Cumulative counts are non-decreasing. Adding this restriction to our optimization would make it even more prone to get stuck in local optima. Therefore, for predicting cumulative deaths, once we get incidence predictions, we convert them to cumulative ones by aggregating the point predictions. We found this gives consistent and stable performance.

### 4.3 Explainability Module

#### Challenges

Policymakers have been constantly placing and lifting travel restrictions and quarantine orders to balance the trade-off between the pandemic burden and economic costs. These policies differ in each administrative region; hence, the data signals that we collect have different meaning and usability at different times and regions. *(C9) Real-time in-sights of forecasts for decision-making and communication:* Decision-makers and public prefer forecasts relatable to epidemiological knowledge, a feature that enables reasoning on their reliability. *(C10) Retrospectively understand signal strengths:* Enabling retrospective analysis of signal strengths over different geographies and periods allow continual improvement of the model.

#### Our approach

See Alg. 2. We selected data ablation accompanied with an interface as means for these two challenges. It is a systematic and simple way to quantify the contribution of signals in the predictions. In Alg. 2, we train models with every group of signals removed and quantify the deviation with respect to a reference point. Then, we rank the groups of signals by this deviation, and check if the contribution of the signal is statistically significant with respect to the model with all signals (this model is denoted as *s* =∅). Our reference can be of two types: (1) available ground truth values for target *y*_*N*+*k*_ until time *N* (lines 8-9), useful to explain strengths of signals in the past; *or* (2) mean of predictions of *s* = ∅ (lines 11-12), useful to enable understanding of the effect of signals in the current predictions. To test *statistical significance*, we use two-sample t-test with null hypothesis *H*_0_ : 𝔼 [*I*(*s*)] = 𝔼 [*I*(∅)] (line 17). This test will tell if removing signals in *s* will truly change predictions. If we fail to reject *H*_0_, then *s* is removed from the ranking output.

##### Interface

To facilitate interactive understanding of the predictions, we constructed a web-based graphical user interface using Vega, a web-based interactive visualization tool. The user interacts with our interface (see Fig. 1) by choosing the region to analyze. The system retrieves a ranking of the signals whose removal impacts the most on the predictions (output of Alg. 2). Then, the user can visualize the predictions with the input predictors for a selected region.

##### Enabled Analysis

Together with Alg. 2 and the interface, our module enables the following two-fold analysis, both driving insights that can inform our data module (feedback loop in Fig. 1). *Real-time insights for (C9:* With our interface, a user can understand which signals are driving the predictive behavior (e.g. trends, slope). In addition, we can reason about the trends in groups of signals and check for group of signals displaying erratic or unreliable behavior. If such signals are found, then we might choose not to include it in our model or we might want to see if there are any issues with the collection, cleaning or transformations (i.e. send them back to the data module). *Retrospective insights for (C10:* Our interface also enables analysis of signal strengths in past predictions. We can evaluate how we could have done in the past given that we had removed some signals (using the training data available in that week). Therefore, we can understand which signals have a positive contribution to our performance and which others have a negative contribution, which ultimately informs our data module.

###### Algorithm 2 Explanations

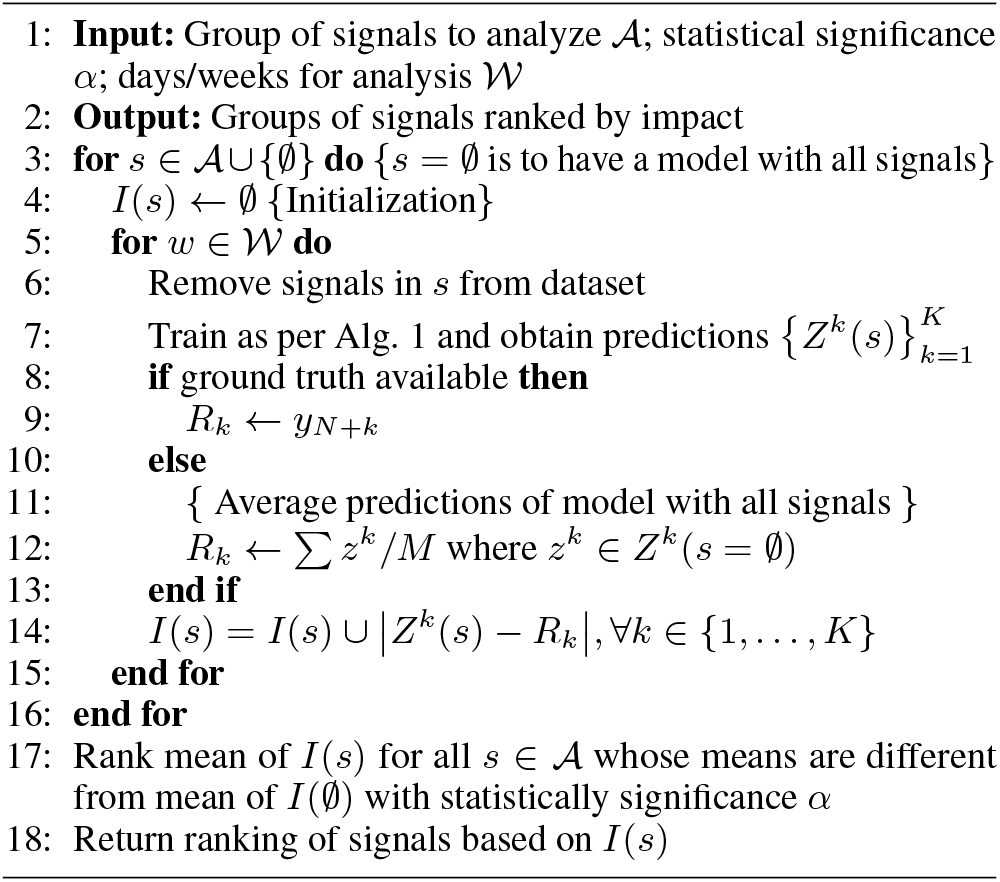

## 5 Empirical Results

We first present the metrics used to evaluate predictive performance, and then make quantitative and qualitative observations about our performance and properties of our forecasts.

### Setup

All experiments are conducted with a Linux machine of 40 processors Intel Xeon CPU E5-2698 v4 @ 2.20GHz, with 252 GB of RAM. Training and obtaining predictions is fast, for a single target takes about 1.5 min. All the results are based on the real-time forecasts submitted during three months (June 8 to September 7 2020).

### Metrics

In epidemic forecasting, predictive performance is usually measured for both point estimates and confidence intervals of our probabilistic distribution of predictions (Tabataba et al. 2017). Hence we utilize the one metric to evaluate each aspect of the predictive power of our method.

For measuring performance of our point estimates, we use the mean absolute percentage error measures the average of absolute percentage error, i.e., 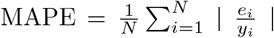 and its value describes how large on average the error is, compared with the actual value.

For measuring performance from a probabilistic perspective, we adopt the probabilistic interval performance metric used in the COVID-19 Forecast Hub introduced in (Bracher et al. 2020). Given the central 1− *α* prediction interval, the interval score Γ_*α*_ is computed as follows:

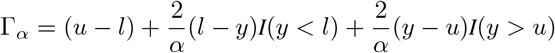

where *y* is ground truth, *I*(·) is the indicator function, *l* corresponds to the 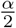 confidence interval number, and *u* corresponds to the 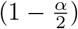 confidence interval number.

### Questions

We aim to demonstrate the our framework Deep-Covid achieves our goals introduced in Section 1. The specific questions we explore via our empirical study are listed below. Recall that the main goal of this paper is to explore the utility of purely data-driven models for emerging pandemics.

Questions Q1-4 directly address this, and Q5 is related to communication, an important asset in a forecasting model.

Q1 Is DeepCovid able to anticipate trend changes? (**G1**: first goal from Section 1, **G2**)

Q2 Does DeepCovid capture finer grain patterns? (**G1, G2**)

Q3 How does DeepCovid perform in US National short-term forecasting? (**G1, G3**)

Q4 Does DeepCovid’s emphasis on short-term forecasting sacrifice longer-term performance? (**G3**)

Q5 Can DeepCovid explain its predictions to epidemiological experts for interpretation? (**G4**)

### 5.1 Observations

#### Observation 1 *[Q1]* DeepCovid *is able to anticipate important changes in trends several weeks ahead*

We are able to anticipate important changes in trends several weeks ahead, which suggests that we achieve goals **G1** and **G2** thanks to our capability of exploiting many heterogeneous signals that are sensitive to what is happening in the ground. Most notably, in Fig. 2(a), we can see we predicted the second peak value and time for US National three weeks early. Our method also predicted with three weeks of anticipation that California, after a stable period, was going to suffer a new increase in deaths (epidemic onset) (Fig. 2(b)).

**Figure 2:**
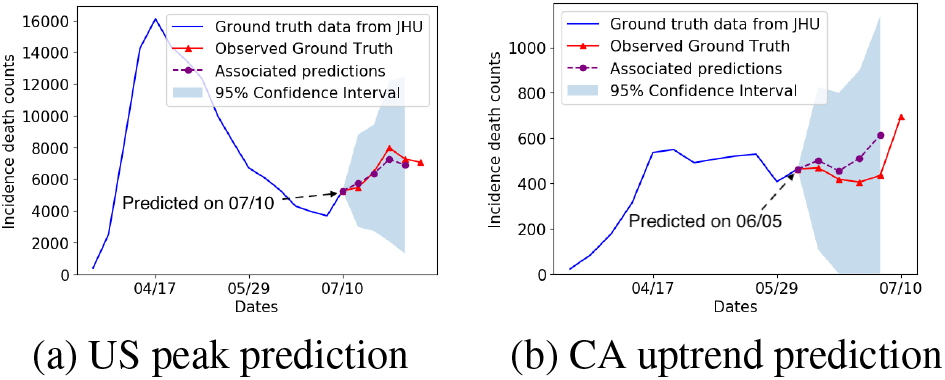
Two examples when we were able to anticipate the upcoming change of trends.

#### Observation 2 *[Q2]* DeepCovid *is able to capture finer grained reporting patterns*

One of the advantages of purely data-driven is that they are able to capture micro patterns in time series data, which brings a complementary forecasting perspective (goals **G1** and **G2**). In forecasting daily hospitalizations, we noticed many regions have the following patterns: P1: drop during weekends; P2: rise on Monday, and continues stable during weekdays. DeepCovid is able to capture them as noted in Fig. 3. In fact, our model is the only approach submitting forecasts to the CDC that captures these patterns^3^.

**Figure 3:**
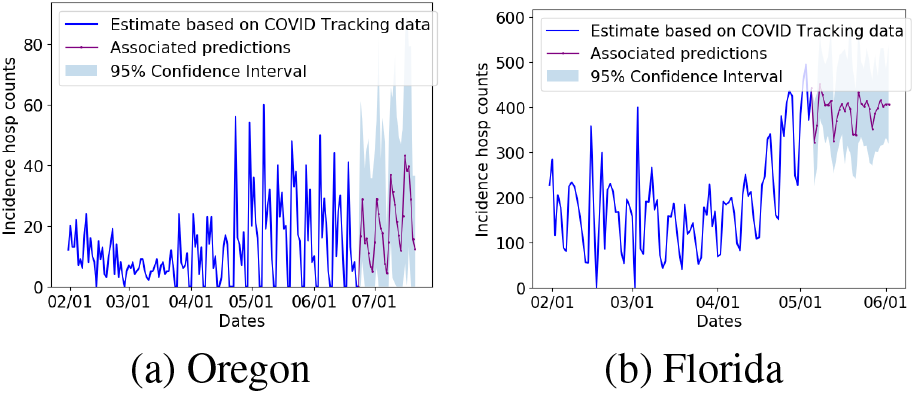
Two examples of finer grained reporting patterns captured by DeepCovid. Note the dips in reporting for weekends.

#### Observation 3 *[Q3]* DeepCovid *excels in US National short-term forecasting*

Here we compare against the official ensemble of all contributing models in the COVID-19 Forecast Hub (including ours). The ensemble has been regarded as one of the best performing models by different independent assessments published on the Web. Needless to say, national-level forecasts are crucial for federal decision makers and are the most visible forecasts in national media. In Fig. 4(a), DeepCovid clearly outperforms this strong baseline in 1- and 2-week ahead across three months. Fig. 4(b) indicates probabilistic performance of our confidence intervals. The fact that we are close suggests we are propagating the uncertainties in the right way. These results show success in goals **G1** and **G3**.

**Figure 4:**
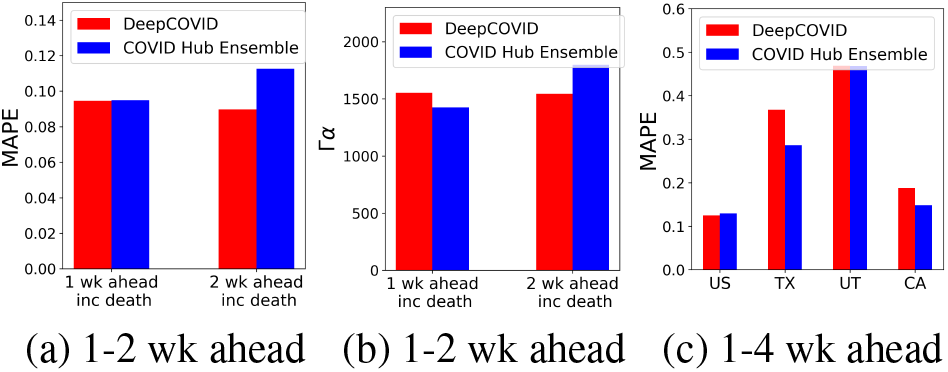
(a) DeepCovid outperforms the official ensemble in US National short-term (1-2 wk ahead) forecasting in MAPE. (b) Our US National short-term confidence intervals are close using probabilistic metric Γ_*α*_ with *α* = 0.7. (c) Our focus on short-term predictions does not compromise longer-term (1-4 wk ahead) performance in multiple regions.

#### Observation 4 *[Q4]* DeepCovid *does not compromise longer-term performance*

In Fig. 4(c), we can notice that our excellence in short-term forecasting does not compromise performance when we consider longer-term predictions in the evaluation. We outperform the ensemble in US National but at state-level our performance is mixed. We are close to this strong baseline in states such as Utah (UT) and California (CA), and slightly farther in Texas (TX). This is indicative that there are still open questions for data-driven forecasting in lower-level granularities, where some signals are more bursty. This is also a good example of where mechanistic models can help. This complements our success for goal **G3**.

#### Observation 5 *[Q5]* DeepCovid *helps explain its predictions to domain experts for interpretation and determine important signals for its predictive performance*

Our explainability module has been key to enable the previously shown quantitative and qualitative performance in real-time forecasting, meeting goal **G4**. This also provides feedback to the data module for selection of signals.

For example, to predict the second US peak in Fig. 2(a), we were able to understand that mobility was the main signal driving this prediction with a clear statistical significance (*α* = 0.05), followed by testing, whose predictions were partially statistically significant. By July, it was unclear if mobility patterns were still capturing social distancing measures, however, the fact that testing signals (which were rapidly increasing in the US) were also contributing to predict the same peak time allowed us to have more confidence in this peak prediction. For predicting the uptrend in Fig. 2(b), using our explainability module, we found that impact of line list, mobility and exposure were statistically significant (with *α* = 0.05) and removing them one at a time did not change the uptrend, which suggests that signals from different groups are being utilized to predict the uptrend. This gave us confidence in our predictions of such important characteristics of the epidemic trajectory.

Similarly, by analyzing over the span of three months, we noticed that the mobility and testing signals were the most important for predictive performance (see Table 3) on average. However, each geographical region may require its own optimized set of data signals as contribution to performance varies by region. For example, including line list had a positive contribution in California, but had a large negative contribution in Texas; this may suggest varying data quality.

**Table 3:** Contribution of signals in 1-4 wk ahead forecasting for US National and three states. We present the t-stat (higher value, higher contribution). Green indicates positive, red negative, and black non-statistically significant contributions.

|  | 1-4 wk ahead |  |  |  |
| --- | --- | --- | --- | --- |
|  | US | CA | GA | TX |
| Line list | 3.51 | 6.09 | 1.34 | -14.17 |
| Mobility | 10.02 | 9.40 | 3.28 | 2.85 |
| Testing | 6.10 | 8.77 | 2.66 | 2.63 |
| Exposure | 3.02 | 1.11 | -0.82 | 0.04 |

## 6 Conclusions

In this paper, we introduced DeepCovid, an operational DL driven framework for real-time COVID forecasting, whose predictions have been submitted to the CDC via COVID-19 Forecast Hub on a weekly basis since April 2020. This was the first purely data driven and DL approach to be submitted to the COVID-19 Forecast Hub. We show that DeepCovid exhibits very good short-term and trend performance, interpretability, principled uncertainty estimation, correlation between forecasts, and ingestion of several data sources despite the novel and fast-moving pandemic scenario which naturally brings several modeling and data challenges. Overall our results give encouraging evidence about using DL for emerging real-time epidemics.

### Discussion

There are several lessons learned from our experience including the observations noted in Section 5. First is the lack of standard data reporting, even among traditional sources (e.g. several states do not report new hospitalizations, or report deaths in different ways causing different lags). This means that while some artifacts can be handled statistically (lags), some mild assumptions are also needed to make meaningful predictions (like for hospitalizations) which are still useful. Second, due to the chaotic situation, a purely data driven model needs to be regularly (weekly) updated to reflect the changing dynamics and data quality, to ensure good performance. Third, explainability is very important, as it can serve as a sanity check based on domain knowledge and also highlight *why* we are making forecasts at different points of the pandemic. Finally, we also found that data revisions are pervasive, and even the *ground truth* can be revised (see appendix), which implies measures of performance may be unreliable till data stabilizes.

Extending our methodology to handle such ‘backfill’ revisions and also work more robustly at smaller scales (e.g. county level), would be interesting. Also, fully automating the deployed version of DeepCovid would be useful e.g. self-discovery of data discrepancies to adapt ‘on-the-fly’. In the prediction module, more end-to-end models for uncertainty estimation should also help in better training and inference. Differentiating between symptomatically similar diseases (such as COVID-19 and flu) would be interesting as well. Finally, we can aim to design more mechanistically motivated neural models to automatically highlight problematic predictions that require further investigation using the explainability module. These directions will enable faster deployment during pandemics.

## Data Availability

All data are publicly available.

https://github.com/CSSEGISandData/COVID-19

https://covidtracking.com/

https://cmu-delphi.github.io/delphi-epidata/

https://www.google.com/covid19/mobility

https://www.apple.com/covid19/mobility

https://gis.cdc.gov/grasp/COVIDNet/COVID19_3.html

https://www.cdc.gov/coronavirus/2019-ncov/covid-data/covidview/04242020/covid-like-illness.html

## Acknowledgments

We would like to thank the anonymous reviewers for their helpful suggestions which improved the paper. This paper is based on work partially supported by the NSF (Expeditions CCF-1918770, CAREER IIS-2028586, RAPID IIS-2027862, Medium IIS-1955883, NRT DGE-1545362), CDC MInD program, ORNL and funds/computing resources from Georgia Tech and GTRI. B. A. was in part supported by the CDC MInD-Healthcare U01CK000531-Supplement.

## Footnotes

1 Indeed, when we started participating, there were only 11 teams in the hub. We were the first two teams predicting hospitalizations.

2 https://www.cdc.gov/coronavirus/2019-ncov/cases-updates/

3 https://projects.fivethirtyeight.com/covid-forecasts/

4 Resources website: http://deepcovid.github.io

5 https://covidtracking.com

6 https://www.hhs.gov/about/agencies/iea/regional-offices/index.html

## References

Adhikari, B., Xu, X., Ramakrishnan, N., and Prakash, B. A. 2019. Epideep: Exploiting embeddings for epidemic forecasting. In Proceedings of the 25th ACM SIGKDD, 577–586.

Altieri, N., et al. 2020. Curating a COVID-19 data repository and forecasting county-level death counts in the United States. arXiv:2005.07882.

Baek, J., et al. 2020. The Limits to Learning an SIR Process: Granular Forecasting for Covid-19. arXiv:2006.06373.

Box, G. E., Jenkins, G. M., Reinsel, G. C., and Ljung, G. M. 2015. Time series analysis: forecasting and control. John Wiley & Sons.

Bracher, J., et al. 2020. Evaluating epidemic forecasts in an interval format. arXiv:2005.12881.

Brooks, L. C., et al. 2018. Nonmechanistic forecasts of seasonal influenza with iterative one-week-ahead distributions. PLOS Computational Biology 14(6).

Chinazzi, M., et al. 2020. The effect of travel restrictions on the spread of the 2019 novel coronavirus (COVID-19) outbreak. Science 368(6489): 395–400.

Dong, E., et al. 2020. An interactive web-based dashboard to track COVID-19 in real time. The Lancet infectious diseases 20(5).

Gelman, A., and Vehtari, A. 2020. What are the most important statistical ideas of the past 50 years? arXiv:2012.00174.

Ginsberg, J., et al. 2009. Detecting influenza epidemics using search engine query data. Nature 457(7232): 1012.

Holmdahl, I., and Buckee, C. 2020. Wrong but Useful — What Covid-19 Epidemiologic Models Can and Cannot Tell Us. New England Journal of Medicine 383(4): 303–305.

Ioffe, S., and Szegedy, C. 2015. Batch Normalization: Accelerating Deep Network Training by Reducing Internal Covariate Shift. volume 37 of Proceedings of Machine Learning Research, 448–456. PMLR.

Jha, A., Ray, S., Seaman, B., and Dhillon, I. S. 2015. Clustering to forecast sparse time-series data. In ICDE, 2015, 1388–1399. IEEE.

Murray, C., et al. 2020. Forecasting the impact of the first wave of the COVID-19 pandemic on hospital demand and deaths for the USA and European Economic Area countries. medRxiv 2020.04.21.20074732.

Osthus, D., et al. 2019. Dynamic Bayesian influenza forecasting in the United States with hierarchical discrepancy (with discussion). Bayesian Analysis 14(1): 261–312.

Ray, E. L., et al. 2020. Ensemble Forecasts of Coronavirus Disease 2019 (COVID-19) in the U.S. medRxiv 2020.08.19.20177493.

Reich, N. G., Niemi, J., House, K., d Hannan, A., Cramer, E., Horstman, S., et al. 2020. covid19-Forecast-Hub: Pre-Publication Snapshot.

Reich, N. G., et al. 2019. A collaborative multiyear, multimodel assessment of seasonal influenza forecasting in the United States. Proceedings of the National Academy of Sciences 201812594.

Shaman, J., Goldstein, E., and Lipsitch, M. 2010. Absolute humidity and pandemic versus epidemic influenza. American journal of epidemiology 173(2): 127–135.

Shaman, J., and Karspeck, A. 2012. Forecasting seasonal outbreaks of influenza. Proceedings of the National Academy of Sciences 109(50): 20425–20430.

Sheridan, C. 2020. Massive data initiatives and AI provide testbed for pandemic forecasting. Nature Biotechnology 38(9): 1010–1013.

Tabataba, F. S., et al. 2017. A framework for evaluating epidemic forecasts. BMC Infectious Diseases 17(1): 345.

Tamerius, J. D., et al. 2013. Environmental predictors of seasonal influenza epidemics across temperate and tropical climates. PLoS pathogens 9(3): e1003194.

Tizzoni, M., et al. 2012. Real-time numerical forecast of global epidemic spreading: case study of 2009 A/H1N1pdm. BMC medicine 10(1): 165.

Venkatramanan, S., et al. 2018. Using data-driven agent-based models for forecasting emerging infectious diseases. Epidemics 22: 43–49.

Volkova, S., Ayton, E., Porterfield, K., and Corley, C. D. 2017. Forecasting influenza-like illness dynamics for military populations using neural networks and social media. PloS one 12(12): e0188941.

Wang, L., Chen, J., and Marathe, M. 2019. DEFSI: Deep learning based epidemic forecasting with synthetic information. In Proceedings of AAAI, volume 33, 9607–9612.

Yuan, Q., et al. 2013. Monitoring influenza epidemics in china with search query from baidu. PloS one 8(5): e64323.

Zhang, Q., et al. 2017. Forecasting seasonal influenza fusing digital indicators and a mechanistic disease model. In Proceedings of WWW, 311–319.

Zou, D., et al. 2020. Epidemic Model Guided Machine Learning for COVID-19 Forecasts in the United States. medRxiv 2020.05.24.20111989.

